# Bridging the gap – estimation of 2022/2023 SARS-CoV-2 healthcare burden in Germany based on multidimensional data from a rapid epidemic panel

**DOI:** 10.1101/2022.12.30.22284061

**Authors:** M Harries, V.K Jaeger, I Rodiah, M.J. Hassenstein, J Ortmann, M Dreier, I von Holt, M Brinkmann, A Dulovic, D Gornyk, O Hovardovska, C Kuczewski, MA Kurosinki, M Schlotz, N Schneiderhan-Marra, M Strengert, G Krause, M Sester, F Klein, A Petersmann, A Karch, B Lange

**Author notes:** indicates shared first authorship (M Harries, V.K Jaeger). indicates shared last authorship (A Karch, B Lange).

## Abstract

Throughout the SARS-CoV-2 pandemic, Germany lacked an adaptive population panel for epidemic diseases and a modelling platform to rapidly incorporate panel estimates.

We evaluated how a cross-sectional analysis of 9922 participants of the MuSPAD study in June/July 2022 combined with a newly developed modelling platform could bridge the gap and analyzed antibody levels, neutralizing serum activity and interferon-gamma release response of serum samples. We categorized the population into four groups with differing protection against severe course of disease (validated by neutralizing serum activity), and found that 30% were in the group with highest protection, and 85% in either the highest categories or second highest group regarding protection level.

Estimated hospitalizations due to SARS-CoV-2 were predicted to be between 30 to 300% of the peak in 02/2021 dependent on assumed variant characteristics. We showed the feasibility of a rapid epidemic panel able to evaluate complex endpoints for SARS-CoV-2 and inform scenario modelling.

## Introduction

During the first two years of the pandemic, Germany lacked rapid adaptive population-based panels for epidemic diseases ^1^ as well as the capacity for central modelling platforms to quickly integrate information from cross-sectional surveys ^2,3,4^. Instead, several population-specific and population-based seroprevalence studies were performed, and the results not published fast enough and rarely used in model estimates or scenarios ^5^. Modelling groups worked largely independently from each other without a central platform for harmonization and integration of results ^6^.

By spring 2022, existing German seroprevalence studies ^2,3,4,7,8,9,10,11,12,13^ had largely ceased sampling and recruiting and had no further funding to conduct new sampling after the Omicron BA.1 and BA.2 waves. Because testing strategies in Germany changed substantially during this time and non-pharmaceutical interventions (NPI) were lifted, the magnitude of underdetection of infections by notified infections to the public health agencies was unclear. This led to modelling efforts with greater uncertainties regarding protection against severe disease or symptomatic infection by vaccination or previous infection for BA.5 and other variants.

Additionally, interpretation of studies on population immunity in terms of their meaning for protection against infection or severe course of disease have become more challenging with larger numbers of re-infections, different vaccination schedules, potentially differential waning immunity, neutralization activity and breakthrough infections ^14^ in different population groups ^15^. Simple seroprevalence surveys are not providing sufficient information, while highly detailed immunological evaluations (i.e. T cells, immune responses towards non-spike antigens) are not scalable to population level studies.

However, estimates indicating protection against severe course of disease are necessary in each modelling study for each new variant using a combination of population-based information on vaccinations, (re)infections confirmed by humoral immunity as well as cellular immunity within various age groups. It is therefore relevant to provide these parameters in a timely fashion, even if they cannot be directly interpreted as protection against infection. Similarly, contact frequency cannot be inferred during a pandemic from previous estimates and should be estimated from current studies ^16^. Using pre-pandemic contact structures for such estimates disregards their profound and differential change in different age groups during a pandemic.

During the past year, we were involved in both, efforts to link new surveys on such complex endpoints of protection against severe disease progression and infection in Germany ^17^ and platforms to harmonize modeling studies that can use the resulting estimates in scenario modeling ^18^. In this process, we transformed an existing “Multilocal and Serial Prevalence Study of Antibodies against SARS Coronavirus 2 in Germany” (MuSPAD) into a rapid, longitudinal, adaptive population-based epidemic panel capable of surveying and sampling within two months of the decision to sample.

We report estimates from this panel on vaccination coverage, re-infection incidence, cellular and humoral immunity as well as contact frequency and intensity, and discuss selected scenarios for the winter 2022/23 in Germany using dynamic models informed by these estimates.

## Methods

### Population-based cohort study

We conducted a new survey of the MuSPAD cohort. The study design has been described previously ^4^. Originally, study participants in MuSPAD were invited in 2020 from a randomly selected list of population registration offices of individuals who lived in the selected study regions. In summer 2022, we amended the study protocol to allow rapid blood sampling and testing for different infectious diseases and resampling of all study participants in the coming years. We now invited all 33,426 original MuSPAD participants from eight study regions by letter or e-mail address to take part in a paper-based or digital survey. 10,090 participants from three study regions (Aachen, Magdeburg and Hannover) were additionally asked to give blood samples onsite. Invitations were reissued by letter to those participants that e-mail addresses were outdated (Figure 1 Flow Chart).

**Figure 1.**
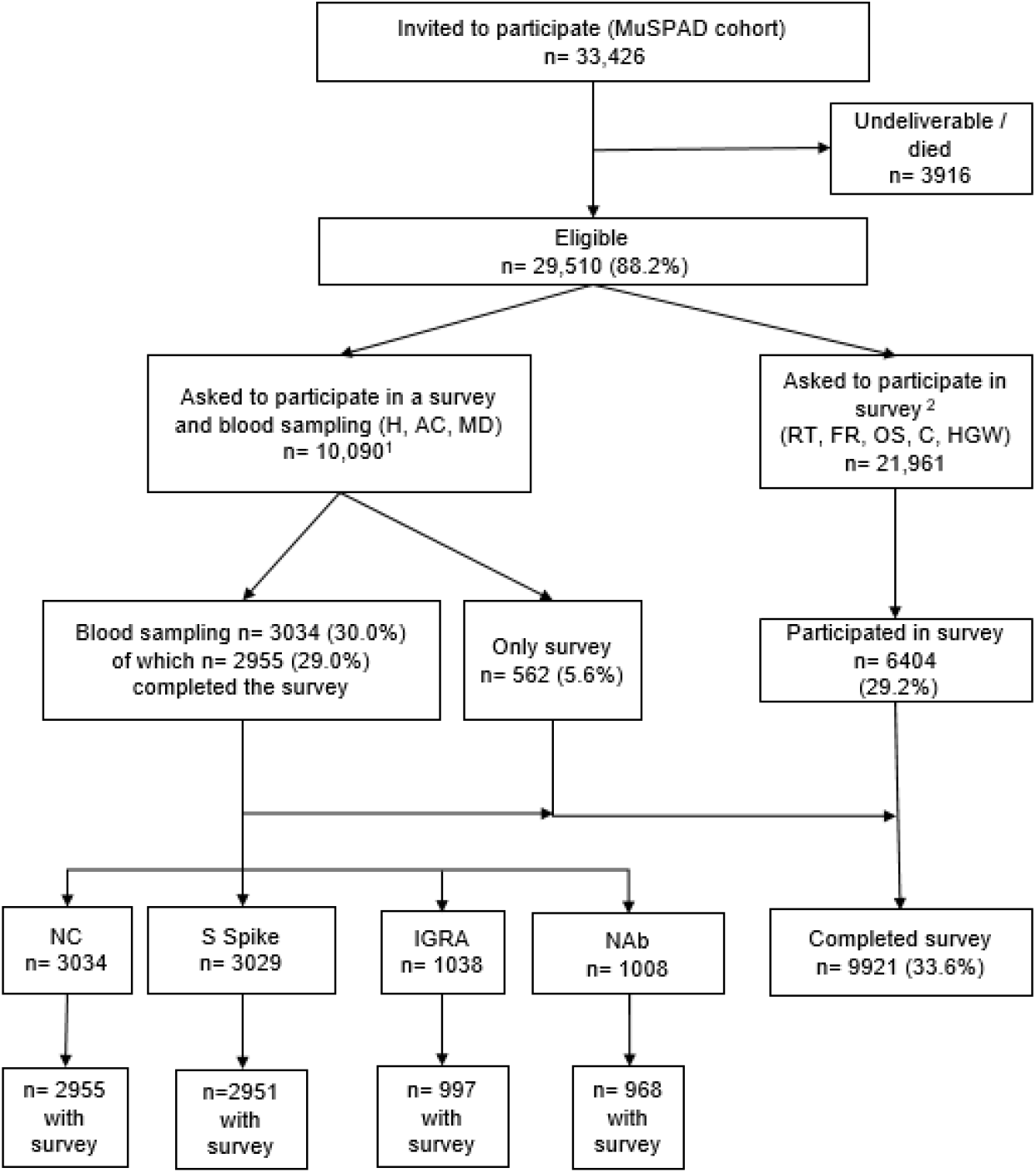
Flow chart of MuSPAD study population and samples analyzed in the seroprevalence study. Samples were tested for IgG antibody against spike protein and nucleoprotein of SARS-CoV-2 (S spike and NC); a subset was confirmed by Interferon-gamma-release Assay (IGRA) and by a Neutralizing Antibody Test (NAb). ^1^ Each blood sampling study site (Hannover (H), Aachen (AC), Magdeburg (MD) had 1,200 appointments available; ^2^ Invited study sites for answering only the questionnaire were Reutlingen (RT), Freiburg (FR), Osnabrueck (OS), Chemnitz (C), Greifswald (HGW).

### Study procedures onsite

Onsite participants gave written informed consent. Blood collection (9-15 ml) was performed by venipuncture using barcoded serum-gel and lithium-heparin monovettes. After centrifugation, the serum-gel monovettes, samples were stored at 4–8°C until analysis. We measured spike S1- and Nucleocapsid (NC) specific IgGs using Elecsys® Anti-SARS-CoV-2 S/N Enzyme-linked Immunosorbent Assays (Roche Diagnostics). Lithium-heparin monovettes were processed according to the manufacturers’ recommendations to determine cell-mediated immunity (CMI). Spike and Non-spike peptides mix targeting full genome (RBD, S1, S2, N, M, NSP) were detected using a QuantiFERON SARS-CoV-2 assay® (Qiagen) ^23^, a SARS-CoV-2 IGRA (interferon gamma releasing assay). Assays were used according to the manufacturer’s instructions. SARS-CoV-2 neutralization potency of sera samples was measured by using lentiviral particles pseudotyped with the spike protein of the Wuhan or the BA.5 isolate, respectively ^24,25^ (Supplement lab analysis).

### Data collection and data management

We developed a questionnaire to update demographic data and health status, as well as new information on SARS-CoV-2 vaccination, known infections and re-infections and previous serologic testing. We ensured that the questionnaire was compatible with a concurrently developed minimal dataset used in the data linkage instrument serohub (www.serohub.net). In this way it could be used within larger projects in Germany, linking data from different population panels within the IMMUNEBRIDGE project ^17,18^.

Data management was conducted according to a data protection concept devised for the original MuSPAD study and approved by relevant authorities including the ethics committee of Hannover Medical School (9086_BO_S_2020). We merged data from the follow-up questionnaires with original MuSPAD data and added new laboratory results.

### Combined endpoints

We performed a targeted literature review ^23,26,27^ focusing on studies with humoral immune markers reflecting protection against infection or hospitalization for Omicron sub variant BA.5 (Supplement Table 1). On this basis, we developed combined endpoints ^18^ to divide the population into groups with different presumptive levels of protection against infection and severe disease progression to parametrize the ordinary differential equation (ODE) model. We divided the population into four main categories based on literature synthesis (Supplement Table 2):

**Table 1.** Characteristics of MuSPAD participants with completed questionnaires in 2022 (overall), with blood samples, and participants with IGRA test results

| Categories | All participants<br>(with or without<br>survey) | Participants<br>with survey<br>and NC results | Participants<br>with survey<br>and S1 results | Participants<br>with survey and<br>IGRA only | Participants<br>with survey and<br>NAb testing |
| --- | --- | --- | --- | --- | --- |
| Total | n=9921 | n=2955 | n=2951 | n=997 | n=968 |
| **Study site** |  |  |  |  |  |
| Aachen | 1040 (10.5%) | 853 (28.9%) | 853 (28.9%) | 104 (10.4%) | 97 (10.0%) |
| Chemnitz | 1356 (13.7%) | 0 (0%) | 0 (0%) | 0 (0%) | 0 (0%) |
| Freiburg | 1498 (15.1%) | 0 (0%) | 0 (0%) | 0 (0%) | 0 (0%) |
| Greifswald | 845 (8.52%) | 0 (0%) | 0 (0%) | 0 (0%) | 0 (0%) |
| Hannover | 1333 (13.4%) | 1103 (37.3%) | 1103 (37.4%) | 0 (0%) | 0 (0%) |
| Magdeburg | 1144 (11.5%) | 999 (33.8%) | 995 (33.7%) | 893 (89.6%) | 871 (90.0%) |
| Osnabrueck | 1515 (15.3%) | 0 (0%) | 0 (0%) | 0 (0%) | 0 (0%) |
| Reutlingen | 1190 (12.0%) | 0 (0%) | 0 (0%) | 0 (0%) | 0 (0%) |
| **Sex** |  |  |  |  |  |
| Diverse | 15 (0.2%) | 1 (0.0%) | 1 (0.0%) | 1 (0.1%) | 1 (0.1%) |
| Female | 5966 (60.6%) | 1774 (60.0%) | 1771 (60.0%) | 637 (63.9%) | 621 (64.2%) |
| Male | 3861 (39.2%) | 1180 (39.9%) | 1179 (40.0%) | 359 (36.0%) | 346 (35.7%) |
| Unknown | 79 (0.8%) | 0 (0%) | 0 (0%) | 0 (0%) | 0 (0%) |
| **Age groups (years)** |  |  |  |  |  |
| 18-34 | 1368 (13.8%) | 280 (9.5%) | 278 (9.4%) | 123 (12.4%) | 118 (12.2%) |
| 35-49 | 1840 (18.7%) | 491 (16.6%) | 491 (16.7%) | 193 (19.4%) | 192 (19.8%) |
| 50-64 | 3331 (33.9%) | 1151 (39.0%) | 1150 (39.0%) | 310 (31.2%) | 301 (31.1%) |
| 65-79 | 2644 (26.9%) | 888 (30.1%) | 887 (30.1%) | 281 (28.2%) | 270 (27.9%) |
| 80+ | 631 (6.4%) | 141 (4.8%) | 141 (4.8%) | 88 (8.8%) | 85 (8.8%) |
| Unknown | 107 (1.1%) | 4 (0.1%) | 4 (0.1%) | 2 (0.2%) | 2 (0.2%) |
| **Education** |  |  |  |  |  |
| Higher education | 5085 (52.7%) | 1729 (59.2%) | 1729 (59.3%) | 517 (55.5%) | 499 (52.3%) |
| Certification after 10 years | 2545 (26.4%) | 681 (23.3%) | 677 (23.2%) | 282 (28.7%) | 274 (28.7%) |
| Certification after 9 years | 1122 (11.6%) | 267 (9.1%) | 267 (9.2%) | 97 (9.9%) | 94 (9.8%) |
| Other Certification | 905 (9.4%) | 245 (8.4%) | 245 (8.4%) | 88 (8.9%) | 88 (9.2%) |
| Unknown | 264 (2.7%) | 33 (1.1%) | 33 (1.1%) | 13 (1.3%) | 13 (1.3%) |
| **Occupation** |  |  |  |  |  |
| Unemployed | 107 (1.1%) | 42 (1.4%) | 42 (1.4%) | 14 (1.4%) | 14 (1.5%) |
| Students | 393 (4.0%) | 82 (2.8%) | 82 (2.8%) | 30 (3.0%) | 29 (3.0%) |
| Teaching sector | 468 (4.8%) | 124 (4.2%) | 123 (4.2%) | 47 (4.7%) | 46 (4.8%) |
| Medical sector | 719 (7.3%) | 190 (6.5%) | 190 (6.5%) | 68 (6.9%) | 67 (7.0%) |
| Retired | 3546 (36.2%) | 1099 (37.4%) | 1099 (37.5%) | 399 (40.2%) | 383 (39.7%) |
| Other | 4558 (46.6%) | 1401 (47.7%) | 1398 (47.6%) | 435 (43.8%) | 425 (43.9%) |
| Unknown | 130 (1.3%) | 17 (0.6%) | 17 (0.6%) | 4 (0.4%) | 4 (0.4%) |
| **Tobacco consumption** |  |  |  |  |  |
| Daily smoker | 1327 (14.4%) | 340 (11.6%) | 339 (11.6%) | 129 (13.1%) | 125 (13.0%) |
| Previous smoker | 3241 (33.2%) | 1032 (35.3%) | 1030 (35.3%) | 322 (32.6%) | 317 (33.1%) |
| Never smoker | 5186 (53.2%) | 1551 (53.1%) | 1550 (53.1%) | 537 (54.4%) | 517 (53.9%) |
| Unknown | 167 (1.7%) | 32 (1.1%) | 32 (1.1%) | 9 (0.9%) | 9 (0.9%) |
| **Household with children under 14 years** |  |  |  |  |  |
| 0 | 6931 (80.5%) | 2019 (82.6%) | 2015 (82.6%) | 670 (81.1%) | 646 (80.8%) |
| 1-2 | 1531 (17.8%) | 401 (16.4%) | 401 (16.4%) | 147 (17.8%) | 146 (18.3%) |
| >2 | 146 (1.7%) | 24 (1.0%) | 24 (1.0%) | 9 (1.1%) | 8 (1.0%) |
| Unknown | 1313 (13.2%) | 511 (17.3%) | 511 (17.3%) | 171 (17.2%) | 168 (17.4%) |
| **Household size** |  |  |  |  |  |
| Lives alone | 2134 (27.5%) | 677 (30.9%) | 676 (30.9%) | 228 (31.4%) | 217 (30.8%) |
| Lives with one to three persons | 4235 (54.5%) | 1204 (55.0%) | 1201 (55.0%) | 400 (55.1%) | 389 (55.3%) |
| Lives with more than three persons | 1397 (18.0%) | 308 (14.1%) | 308 (14.1%) | 98 (13.5%) | 98 (13.9%) |
| Unknown | 2155 (21.7%) | 766 (25.9%) | 766 (26.0%) | 271 (27.2%) | 264 (27.3%) |
| **Number of COVID vaccine doses** |  |  |  |  |  |
| None | 308 (3.26%) | 57 (2.0%) | 57 (2.0%) | 34 (3.5%) | 34 (3.6%) |
| One dose | 70 (0.741%) | 20 (0.7%) | 20 (0.7%) | 6 (0.6%) | 6 (0.6%) |
| Two doses | 592 (6.3%) | 144 (4.96%) | 143 (4.9%) | 68 (7.0%) | 65 (6.8%) |
| Three doses | 6982 (73.9%) | 2179 (75.0%) | 2176 (75.0%) | 751 (76.7%) | 730 (76.8%) |
| Four doses | 1491 (15.8%) | 505 (17.4%) | 505 (17.1%) | 120 (12.3%) | 115 (11.9%) |
| Unknown | 478 (4.8%) | 50 (1.7%) | 50 (1.7%) | 18 (1.8%) | 18 (1.9%) |
| **Comorbidities** |  |  |  |  |  |
| Hypertension | 2834 (28.8%) | 929 (31.4%) | 928 (31.4%) | 348 (34.9%) | 333 (34.4%) |
| No hypertension disease | 7009 (71.2%) | 2026 (68.6%) | 2023 (68.6%) | 649 (65.1%) | 635 (65.6%) |
| Unknown | 78 (0.8%) | 0 (0%) | 0 (0%) | 0 (0%) | 0 (0%) |
| Diabetes mellitus | 610 (6.2%) | 178 (6.0%) | 177 (6.0%) | 83 (8.3%) | 82 (8.5%) |
| No diabetes mellitus | 9233 (93.8%) | 2777 (94.0%) | 2774 (94.0%) | 914 (91.7%) | 886 (91.5%) |
| Unknown | 78 (0.8%) | 0 (0%) | 0 (0%) | 0 (0%) | 0 (0%) |
| Cardiovascular | 923 (9.4%) | 292 (9.9%) | 292 (9.9%) | 115 (11.5%) | 113 (11.7%) |
| No cardiovascular disease | 8920 (90.6%) | 2663 (90.1%) | 2659 (90.1%) | 882 (88.5%) | 855 (88.3%) |
| Unknown | 78 (0.8%) | 0 (0%) | 0 (0%) | 0 (0%) | 0 (0%) |
| Chronic lung disease | 754 (7.7%) | 230 (7.8%) | 230 (7.8%) | 94 (9.4%) | 92 (9.5%) |
| No chronic lung disease | 9089 (92.3%) | 2725 (92.2%) | 2721 (92.2%) | 903 (90.6%) | 876 (90.5%) |
| Unknown | 78 (0.8%) | 0 (0%) | 0 (0%) | 0 (0%) | 0 (0%) |
| Immunosuppression disease | 228 (2.3%) | 65 (2.2%) | 65 (2.2%) | 20 (2.0%) | 18 (1.9%) |
| No immunosup-<br>pression disease | 9615 (97.7%) | 2890 (97.8%) | 2886 (97.9%) | 977 (98.0%) | 950 (98.1%) |
| Unknown | 78 (0.8%) | 0 (0%) | 0 (0%) | 0 (0%) | 0 (0%) |
| Cancer | 218 (2.2%) | 61 (2.1%) | 61 (2.1%) | 17 (1.7%) | 18 (1.9%) |
| No cancer | 9625 (97.8%) | 2894 (97.9%) | 2890 (97.9%) | 980 (98.3%) | 950 (98.1%) |
| Unknown | 78 (0.8%) | 0 (0%) | 0 (0%) | 0 (0%) | 0 (0%) |
| Post COVID | 121 (1.2%) | 28 (0.9%) | 28 (0.9%) | 8 (0.8%) | 8 (0.8%) |
| No Post COVID | 9722 (98.8%) | 2927 (99.1%) | 2923 (99.1%) | 989 (99.2%) | 960 (99.2%) |
| Unknown | 78 (0.8%) | 0 (0%) | 0 (0%) | 0 (0%) | 0 (0%) |
| **Number of comorbidities** |  |  |  |  |  |
| 0 | 5984 (60.3%) | 1686 (57.1%) | 1683 (57.0%) | 531 (53.3%) | 515 (53.2%) |
| 1 | 2687 (27.1%) | 878 (29.7%) | 878 (29.8%) | 299 (30.0%) | 293 (30.3%) |
| 2-3 | 1191 (12.0%) | 380 (12.9%) | 379 (12.8%) | 162 (16.2%) | 155 (16.0%) |
| >3 | 59 (0.6%) | 11 (0.4%) | 11 (0.4%) | 5 (0.5%) | 5 (0.5%) |
| **Received flu vaccine 2021/2022** |  |  |  |  |  |
| Yes | 4818 (48.9%) | 1767 (59.8%) | 1766 (59.8%) | 640 (64.2%) | 617 (63.7%) |
| No | 5025 (51.1%) | 1188 (40.2%) | 1185 (40.2%) | 357 (35.8%) | 351 (36.3%) |
| Missing | 78 (0.8%) | 0 (0%) | 0 (0%) | 0 (0%) | 0 (0%) |
| **COVID vaccination in fall 2022** |  |  |  |  |  |
| Yes | 9148 (96.5%) | 2852 (96.5%) | 2848 (97.9%) | 947 (96.3%) | 919 (96.2%) |
| No | 308 (3.3%) | 57 (1.9%) | 57 (2.0%) | 34 (3.5%) | 34 (3.6%) |
| I don't know /<br>want to answer | 22 (0.2%) | 4 (0.1%) | 4 (0.1%) | 2 (0.2%) | 2 (0.2%) |
| Missing | 443 (4.5%) | 42 (1.4%) | 42 (1.4%) | 14 (1.4%) | 13 (1.3%) |

**Table 2.**
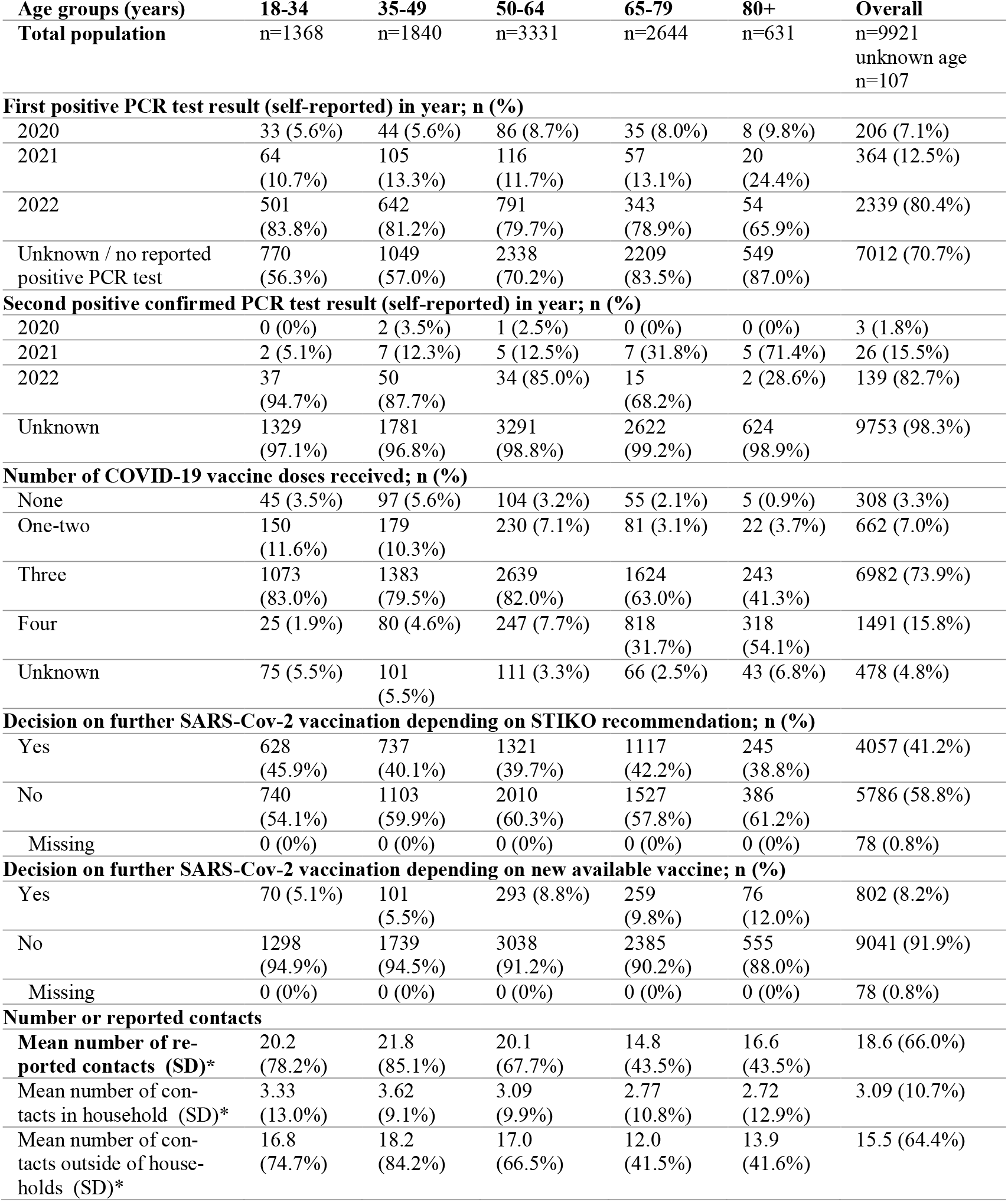
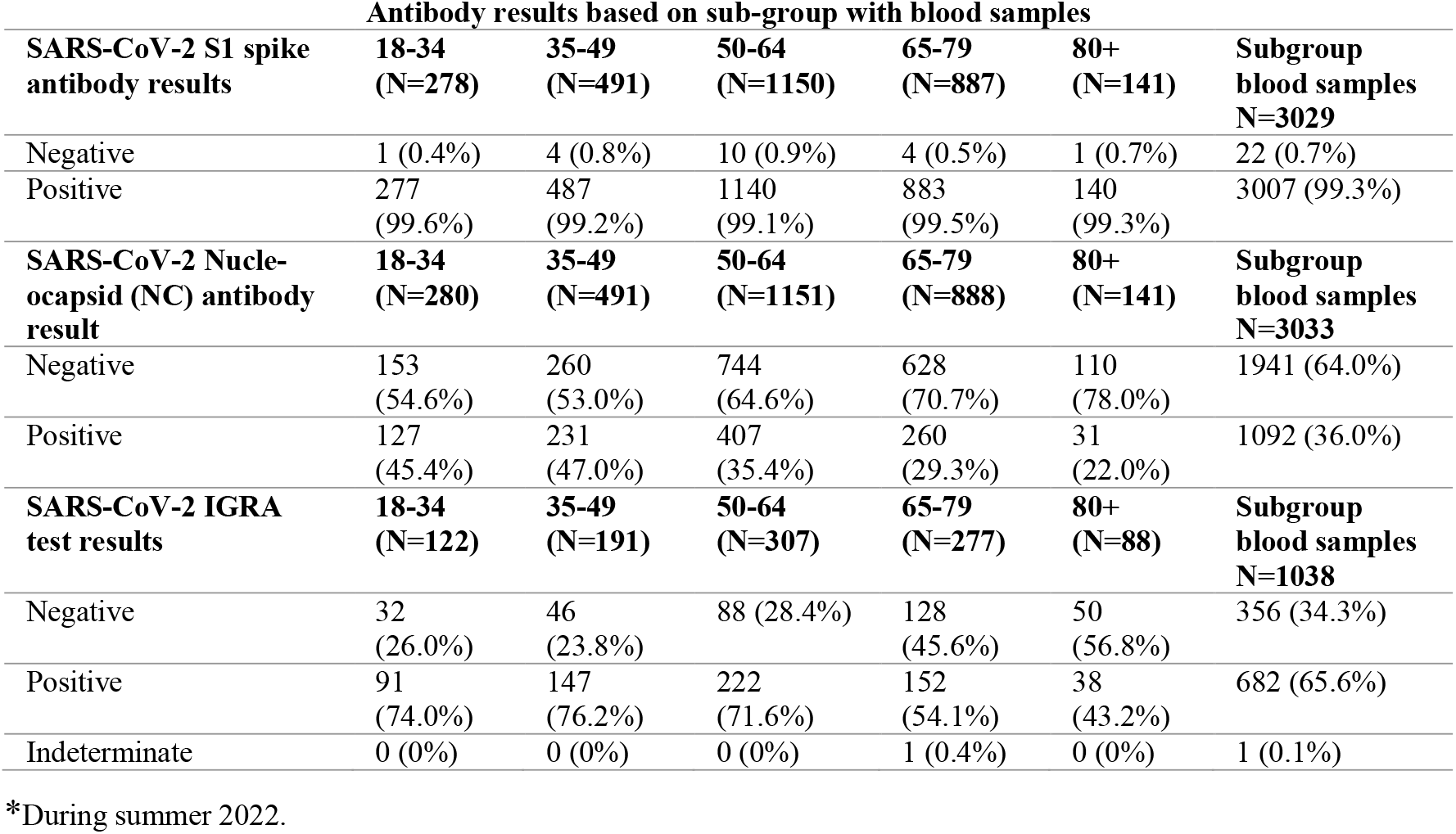
Self-reported SARS-CoV-2 positive test history, vaccination status, confirmed humoral or cellular immunity test result (S- and NC-antibodies, IGRA) and number of social contacts for defined age groups of MuSPAD participants by age group.

Group I has a likely high level of protection against severe course of disease based on four exposures (infection and/or vaccination) with one of them occurring in 2022 confirmed by humoral immune correlates. Group Ia in contrast to group Ib has in addition a positive cell-mediated immune (CMI) response.

Group II was defined as having a moderate protection against severe course of disease based on three exposures with humoral immune correlates. Group IIa has in contrast to group IIb in addition a positive CMI response.

Group III has a low level of protection against severe course of disease based on three exposures without immune correlate, or 1-2 exposures with or without immune correlate or 0 exposures with immune correlate. Group IIIa has in contrast to group IIIb in addition a positive CMI response.

Group IV has no protection against severe course of disease with no reported exposures and undetected cellular and humoral responses.

### Data analysis

We present sociodemographic data, current vaccination coverage, history of infections and reinfections as well as humoral and cellular immunity characteristics by standard descriptive statistics. To evaluate individual and aggregate changes in prevalence and titer of IgG antibodies against spike and nucleocapsid over time we compared results from before 2022 analysed with the MULTICOV-AB assay, a multiplex assay based on the Luminex platform ^28^, with current results.

The four combined endpoints were described for adult age groups, sex and underlying conditions. We summarized contact behavior and infection rate over time, for age groups and vaccination status. We conducted random-effect logistic and binomial regression models on the determinants vaccination coverage, re-infections and humoral and cellular immunity as well as exposure status. Analyses were performed using R Version 4.0.2.

### Age-specific SEIR model

We applied a deterministic SEIR (Susceptible-Exposed-Infectious-Recovered) model with a realistic age structure and contact behavior based on a social contact matrix ^16^. The model has been previously described ^22^. We included compartments for hospitalizations, patients in the intensive care unit (ICU), and deaths. In one version of this model, we assumed that the susceptible population is split into four groups in accordance with the combined endpoint results (Supplement Table 3) based on humoral confirmation; in a third version of the model we similarly split the susceptible into four groups but additionally divided according to IGRA (Interferon-Gamma-Release-Assay) positivity. Figure 2 shows the structure of these models. In addition to the established compartments, we added estimates of confirmed exposure from the above-mentioned population-based study in each age group and parametrized according to the literature review performed. Figure 2 also provides an overview of the scenarios that we designed: Scenario A1 stimulates a wave of a BA.5-like variant without (booster) vaccinations; Scenario A2 covers a BA.5-like variant with a booster campaign using an adapted vaccine. Scenario B models a new variant with BA.5-like capacities but higher transmissibility without (B1) and with a variant-adapted vaccine booster campaign (B2). Scenario C models a new variant with higher severity and transmissibility without (C1) and with a booster campaign (C2). Scenario D models a new variant with higher transmissibility, severity and immune evasion without (D1) and with a booster campaign (D2). Scenario E models a new variant with even higher transmissibility and severity and immune evasion without (E1) and with a booster campaign (E2).

**Table 3.** Random effects logistic regression analysis of participant’s characteristics on subgroups according to different endpoints and laboratory results (IgG antibodies and IGRAs)

|  | Odds of having highest combined endpoint vs. all others (n=2640) |  |  | Odds of having highest two endpoints and pos. IGRA vs. all others (n=808) |  |  | Odds of having pos. IGRA vs. negative (n=815) |  |  |
| --- | --- | --- | --- | --- | --- | --- | --- | --- | --- |
| Age groups (years) | OR | 95% CI | p-value | OR | 95% CI | p-value | OR | 95% CI | p-value |
| 18-24 (reference) | 1 | - | - | 1 | - | - | 1 | - | - |
| 35-49 | 1.12 | 0.79 – 1.59 | 0.540 | 1.12 | 0.62 – 2.00 | 0.713 | 1.16 | 0.63 – 2.14 | 0.641 |
| 50-64 | 0.76 | 0.55 – 1.05 | 0.095 | 0.54 | 0.30 – 0.98 | 0.044 | 0.92 | 0.53 – 1.63 | 0.787 |
| 65-79 | 1.62 | 1.14 – 2.30 | 0.007 | 1.03 | 0.56 – 1.90 | 0.926 | 0.40 | 0.22 – 0.72 | 0.002 |
| 80+ | 3.34 | 1.92 – 5.80 | <0.001 | 0.83 | 0.35 – 1.93 | 0.658 | 0.30 | 0.14 – 0.65 | 0.002 |
| Sex |  |  |  |  |  |  |  |  |  |
| Females (reference) | 1 | - | - | 1 | - | - | 1 | - | - |
| Males | 0.98 | 0.81 – 1.18 | 0.808 | 0.90 | 0.63 – 1.30 | 0.575 | 0.91 | 0.65 – 1.28 | 0.597 |
| Employment & education |  |  |  |  |  |  |  |  |  |
| Employed in teaching sector | 1.75 | 1.14 – 2.68 | 0.010 | 1.07 | 0.48 – 2.40 | 0.867 | 0.56 | 0.27 – 1.16 | 0.119 |
| Employed in medical sector | 1.98 | 1.40 – 2.79 | <0.001 | 1.52 | 0.80 – 2.87 | 0.197 | 0.84 | 0.43 – 1.66 | 0.615 |
| Employed in another field (reference) | 1 | - | - | 1 | - | - | 1 | - | - |
| Higher education certificate | 0.89 | 0.74 – 1.07 | 0.218 | 0.97 | 0.68 – 1.39 | 0.883 | 1.09 | 0.78 – 1.53 | 0.594 |
| No education (reference) | 1 | - | - | 1 | - | - | 1 | - | - |
| Smoking |  |  |  |  |  |  |  |  |  |
| Never (reference) | 1 | - | - | 1 | - | - | 1 | - | - |
| Current | 0.88 | 0.65 – 1.18 | 0.389 | 1.11 | 0.65 – 1.89 | 0.705 | 2.30 | 1.32 – 4.01 | **0.003** |
| Former | 1.07 | 0.88 – 1.30 | 0.498 | 1.15 | 0.79 – 1.68 | 0.458 | 1.20 | 0.84 – 1.72 | 0.319 |
| Kids aged <= 14 within household |  |  |  |  |  |  |  |  |  |
| None (reference) | 1 | - | - | 1 | - | - | 1 | - | - |
| 1 or 2 | 1.47 | 1.12 – 1.92 | **0.005** | 1.52 | 0.91 – 2.55 | 0.114 | 1.64 | 0.93 – 2.92 | 0.090 |
| 3 and more | 0.66 | 0.25 – 1.75 | 0.404 | 1.61 | 0.36 – 7.23 | 0.535 | 0.22 | 0.05 – 0.92 | 0.039 |
| Comorbidities (reference: absence of respective comorbidity) |  |  |  |  |  |  |  |  |  |
| hypertension | 1.16 | 0.94 – 1.42 | 0.164 | 1.48 | 0.99 – 2.21 | 0.058 | 1.51 | 1.04 – 2.20 | **0.032** |
| diabetes | 1.41 | 0.96 – 2.08 | 0.081 | 1.02 | 0.51 – 2.04 | 0.947 | 0.75 | 0.39 – 1.43 | 0.385 |
| cardiovascular | 1.46 | 1.08 – 1.97 | **0.013** | 1.57 | 0.92 – 2.68 | 0.096 | 1.25 | 0.74 – 2.12 | 0.410 |
| lung disease | 1.30 | 0.94 – 1.78 | 0.113 | 0.76 | 0.41 – 1.38 | 0.364 | 0.45 | 0.27 – 0.78 | **0.004** |
| immunosuppression | 1.82 | 1.03 – 3.23 | **0.040** | 2.00 | 0.69 – 5.74 | 0.200 | 0.77 | 0.26 – 2.29 | 0.644 |
| cancer | 2.40 | 1.28 – 4.51 | **0.007** | 1.24 | 0.30 – 5.09 | 0.769 | 1.51 | 1.04 – 2.20 | **0.032** |

Note: Study location constituted the random effects. Seven participants had no combined endpoint.

**Figure 2.**
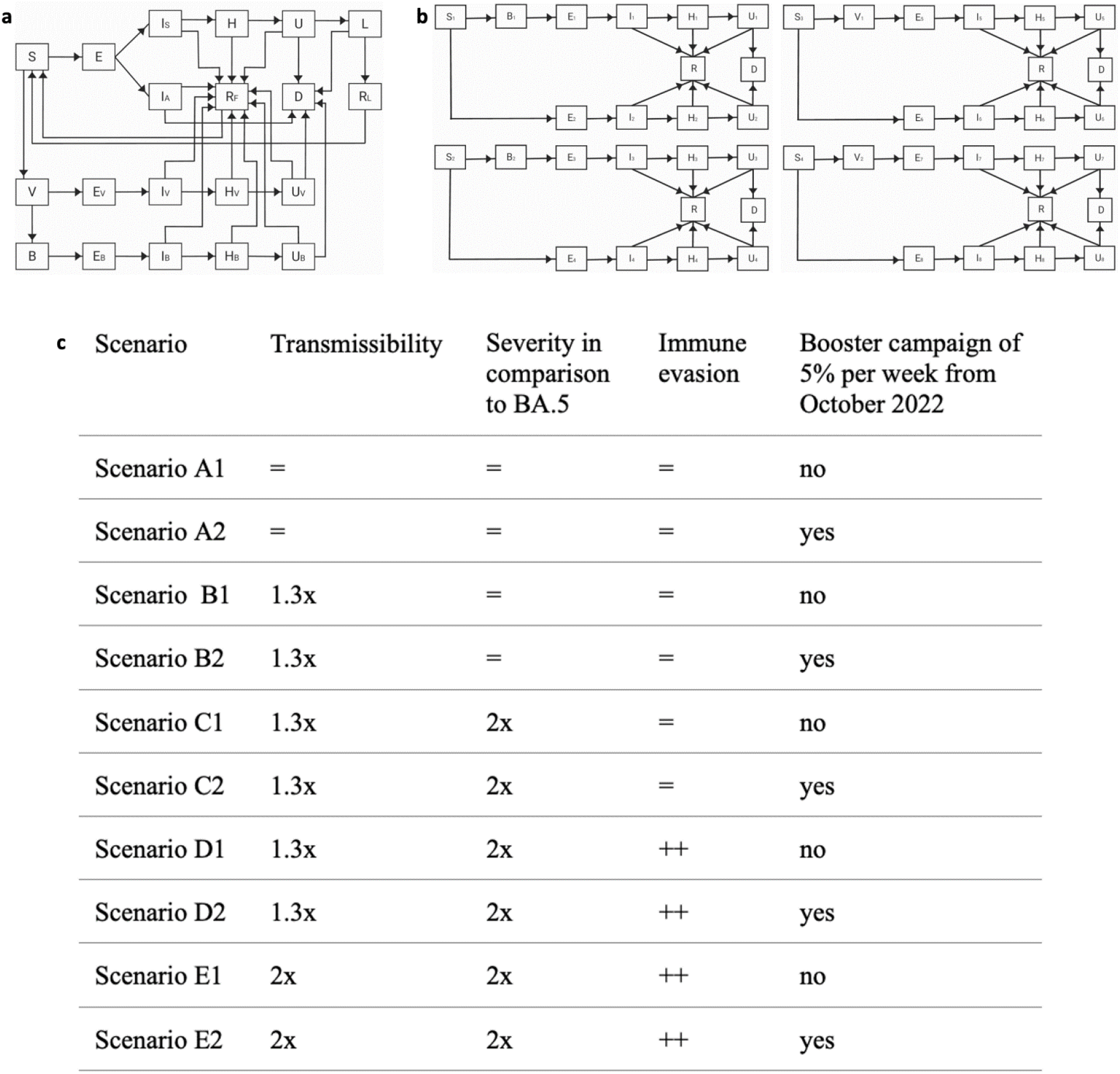
**a:** Structure of the models based on officially recorded public health data. The compartments are classified either as susceptible (S), exposed (E), asymptomatically infectious (I_A_), symptomatically infectious (I_S_), hospitalized (H), in intensive care (U), suffering under long-COVID (L), fully recovered (R_F_), recovered from long-COVID (R_L_), dead (D), vaccinated (V), exposed after vaccination (E_V_), infectious after vaccination (I_V_), hospitalized after vaccination (H_V_), in intensive care after vaccination (U_V_), booster (B), exposed after booster (E_B_), infectious after booster (I_B_), hospitalized after booster (H_B_), and in intensive care after booster (U_B_). **b:** Structure of the two models based on estimates from the population-based panel with humoral and with cellular immunity. The compartments are classified either as susceptible (S), exposed (E), infectious (I), hospitalized (H), in intensive care (U), recovered (R), dead (D), vaccinated (V), and booster (B). **c:** Designed scenarios to model the pandemic course for winter 2022/23 in Germany. Transmissibility is estimated by the basic reproduction number.

For the description of the population-based cohort, we followed STROBE ^19^ and adapted the WHO protocol for SARS-CoV-2 seroprevalence studies ^20^. We used EPIFORGE ^21,22^ to describe the ODE model.

## Results

In June 2022, we re-invited all previous MuSPAD participants (n=33,426) from eight regions in Germany (Figure 1) to take part in a survey, of whom 9922 participated (30%). The majority of participants answered questionnaires within six days (range 1-122 days). Among 10,090 MuSPAD participants from three regions (Aachen, Hannover, Magdeburg) we collected 3034 blood samples, of which 2955 completed the questionnaire (Figure 1). In a subgroup of 1038 individuals from two centers (Aachen, Magdeburg), additionally IGRAs were performed, and in 1008 individuals we tested for SARS-CoV-2 neutralizing activity of serum IgG.

### Characteristics of the study population

More women than men (60.1% vs 39.2%) took part in the survey, the mean age was 54.8 years (IQR 44-68 years) and 52.7% had a higher education certificate (Table 1). Diabetes prevalence was 6.2%, 28.8% of the participants had hypertension and twelve percent described two or more predisposing diseases. Seven percent were employed in a hospital or clinic, 4.8% in the school sector, 36.2% were retired and 46.6% working in other fields. Nearly twenty percent of the participants had children under the age of 14 living in their household.

### Population estimates of vaccination coverage, re-infections, humoral and cellular immunity, and contact frequency

In Table 2 we present population estimates of vaccination coverage, (re)-infections, seropositivity for NC and S antibodies and SARS-CoV-2 IGRA positivity for different age groups. Of 9,921 participants who answered the survey, more than 85% received at least three doses of a SARS-CoV-2 vaccine. Of those with a reported positive PCR test (29.3%), seven percent (7.1%) reported having had a confirmed positive PCR test in 2020, 12.5% in 2021 and 80.4% in 2022. Of those with self-reported reinfection (1.7%), this occurred in 2020 in three (1.8%) participants, in twenty-six (15.5%) in 2021 and in 139 (82.7%) in 2022. Mean number of contacts reported in summer 2022 was 3.1 inside the household and 15.5 outside of the household (Table 2).

Of the 3033 participants with blood samples, 99.3% had antibodies against the S-antigen and 36.0% had antibodies against the NC-antigen during sampling between June and July 2022. Of the 1038 participants with IGRA results, 65.6% were IGRA positive and one participant had an indeterminate result (0.1%). IGRA-positivity was lower in those > 65 years of age (Table 2).

### Confirmed exposure to SARS-CoV-2 infection or vaccination

When categorizing the population into four groups according to exposure and humoral immunity, we found that 34.2% reported four exposures (including one in 2022) confirmed by humoral immunity. This proportion was higher in those above 65 years. More than 95.4% reported at least three exposures confirmed by humoral immunity. Of those with available test who reported at least four exposures with one in 2022 confirmed by humoral immune correlates, 24.5% did not show a positive IGRA result, this proportion increased with age (Supplement Table 2, Figure 3).

**Figure 3.**
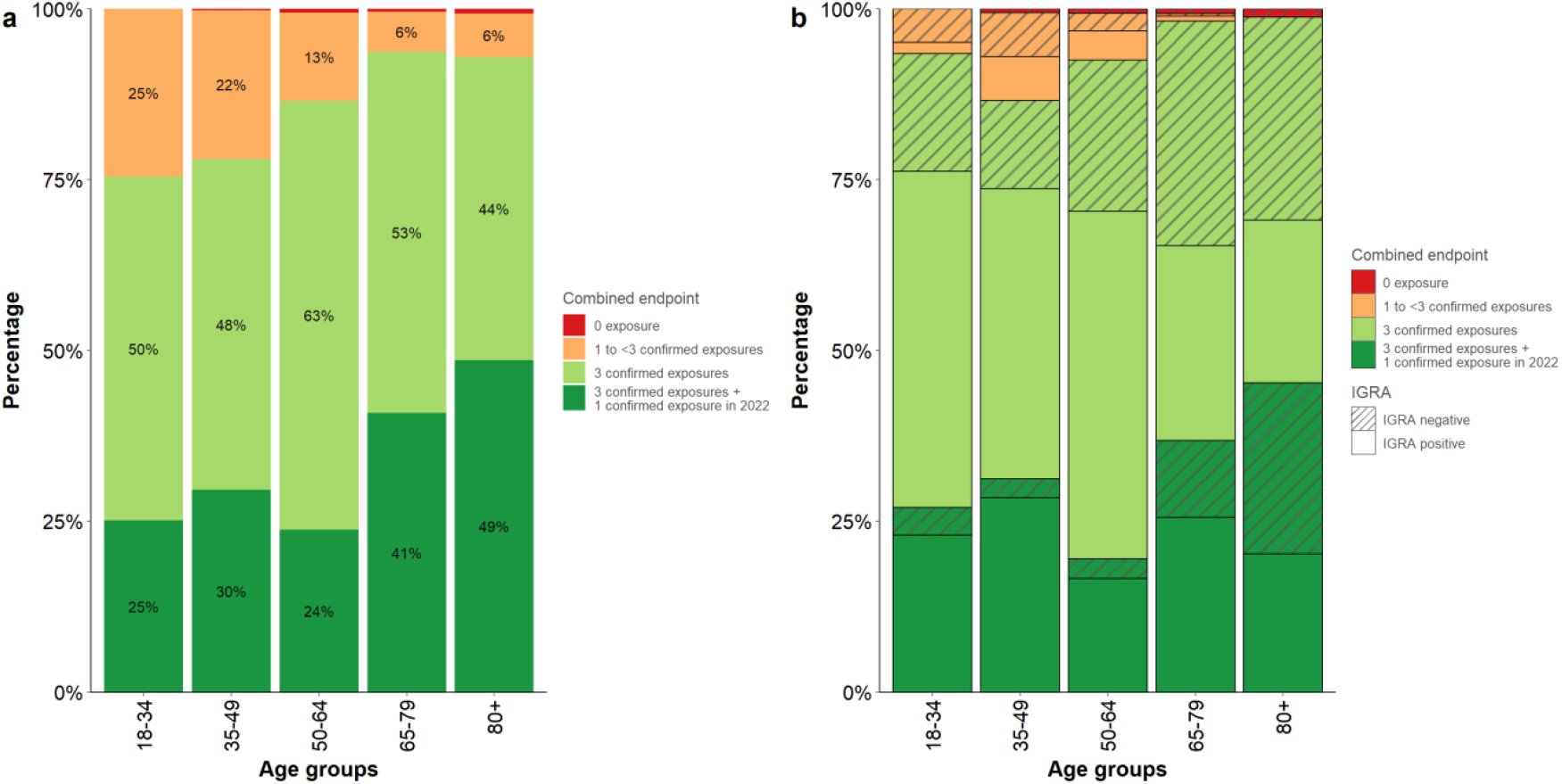
Combined endpoint representing exposures by infection or vaccination with corresponding humoral immune response stratified by age (**a**) and further stratified by IGRA positivity (**b**). Note: **(a)** is based on all participants with information on the combined endpoint and age (n=3209) and **(b)** is based on all participants with information on the combined endpoint, age and IGRA (n=974).

Those with blood samples and aged over 80 years had three times the odds of having four exposures confirmed by humoral immune correlates (OR 3.34; 95% CI 1.92-5.80). Employment in the medical field or the education sector and having 1-2 children was also associated with having higher odds of having had four exposures, respectively (Table 3).

Having at least three exposures and a positive IGRA was not associated with older age (OR 0.83; 95% CI 0.35-1.93). Those aged 80 or older had lower odds of having a positive IGRA (OR 0.30; 95% CI 0.14-0.65). Having a chronic lung disease was associated with lower odds of having a positive IGRA (OR 0.45; 95% CI 0.27-0.78) while current smoking was associated with higher odds for IGRA positivity (OR 2.30; 95% CI 1.32-4.01) (Table 3).

### Correlation of neutralization vs BA.5 to exposure endpoint

The neutralizing SARS-CoV-2 antibody response (NAb) was higher in participants with three or more exposures confirmed by humoral response. Participants showed higher NAb activity against parental Wu01 variant compared to BA.5 variant (Figure 4). The majority of individuals had high S-reactive IgG levels, which correlated with the number of exposures. Anti-NC antibody titers were more scattered across the combination endpoints (Supplement Figure 1).

**Figure 4.**
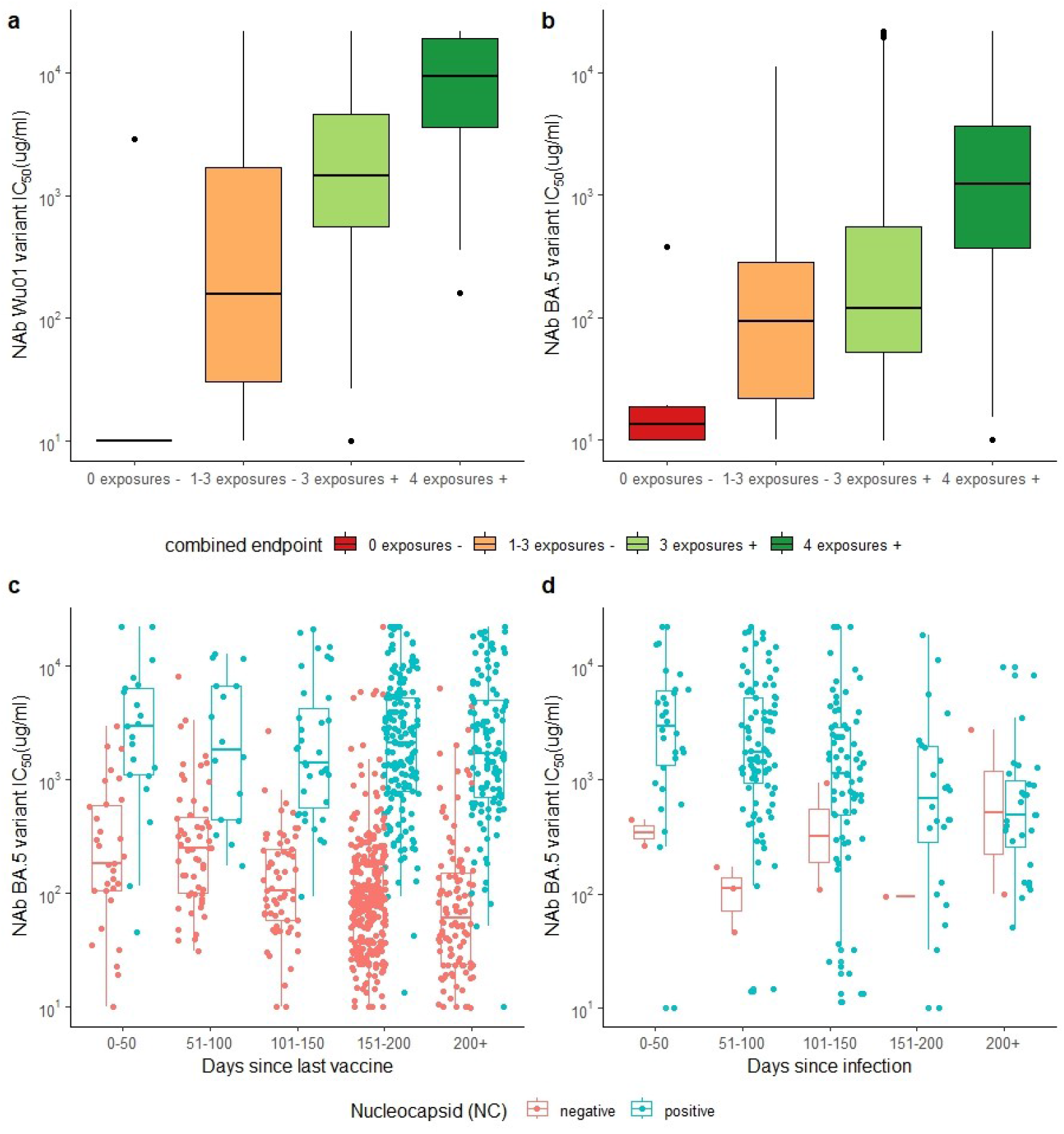
MuSPAD participants neutralizing antibody responses to variants BA.5 (**a**) and Wu01 (**b**) over combined endpoints in 2022; a boxplot of BA.5 neutralizing antibody response by time since last SARS CoV-2 vaccination (**c**) and last SARS-CoV-2 infection (**d**) stratified by nucleocapsid antibody response (NC with ≥0.8 BAU/ml cut-offs for seropositivity).Combined endpoints: 4 exposures +: 4 exposures (vaccination or infection) with humoral immune correlates including one infection/vaccination in 2022; 3 exposures +:3 exposures (vaccination or infection) with humoral immune correlates; 1-3 exposures: 1-3 exposures with or without immune correlate or 0 exposures with at least 1 positive humoral immune correlate; 0 exposure:

Previous infection confirmed by NC antibodies modified the decreasing trend in neutralizing antibody titers against BA.5 and Wu01 with increasing time since last vaccination seen in those without NC antibodies. In contrast to participants without prior infection, there was no decreasing trend in participants with NC antibodies. Participants with more than 150 days since the last vaccination had lower neutralization activity than participants with vaccinations in the last 50 days in the absence of NNC-antibodies. Participants with confirmed infection (positive NC antibodies) showed a clear downward trend of neutralization activity over time (Figure 4). This trend was not as clear for the interferon-gamma-release (Supplement Figure 2).

### Vaccinations, (re)infections, contact frequency and immunity markers over time

MuSPAD provided measurements at six time points during the pandemic, in September 2020, in November 2020, in February 2021, in May 2021, in August 2021 and in June 2022. In Figure 5 we show aggregate summary estimates of seropositivity (Figure 5a) over the measurement time points (5a), estimated proportions of those ever infected with SARS-CoV-2 according to N-antibody positivity (Figure 5b), proportions of self-reported infections and reinfections according to reported dates (Figure 5c), and proportions of individuals with first, second, third, and fourth vaccine doses (Figure 5d) for participants included from these time points. In Figure 5e, we show the contact frequency over time.

**Figure 5.**
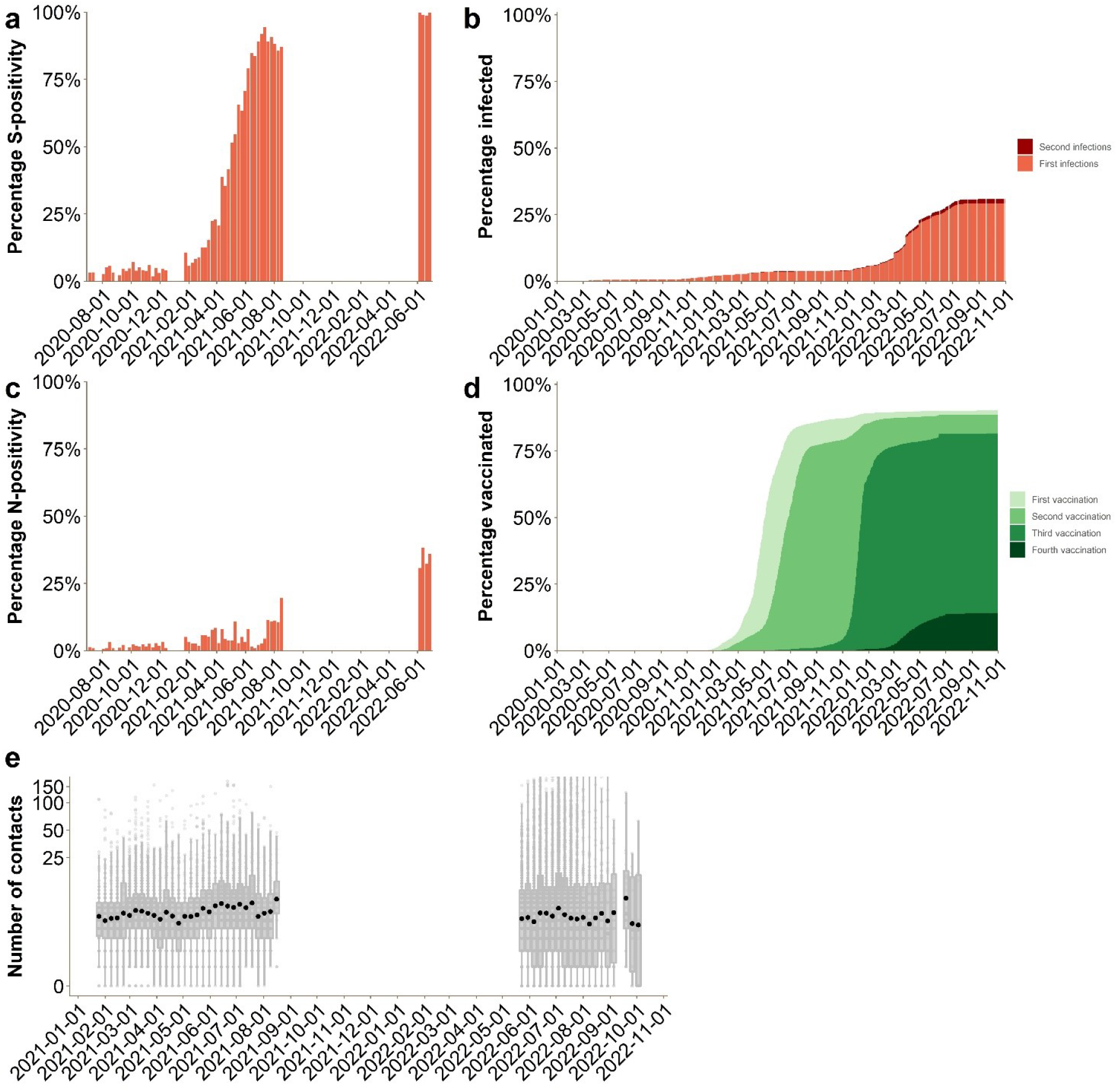
Aggregated estimates of SARS-CoV-2 seropositivity (**a**), estimated proportion of those ever infected with SARS-CoV-2 according to N-antibody positivity (first infection) (**b**), proportion of self-reported infections and reinfections by time (second infection) (**c**), proportion of individuals with first, second, third and fourth vaccine doses (**d**) over time and self-reported contact frequencies over time (**e**; sampling/survey break during September 2021 and May 2022).

### Potential healthcare burden during winter 2022/23 in Germany

We looked at five scenarios with and without booster campaigns and described peak general ward hospitalizations and peak ICU hospitalizations (Figure 2c). Figures 6A to E show how infections, hospitalizations (Supplement Figure 3) and ICU hospitalizations have been modelled in each scenario for three different compartmental models.

**Figure 6.**
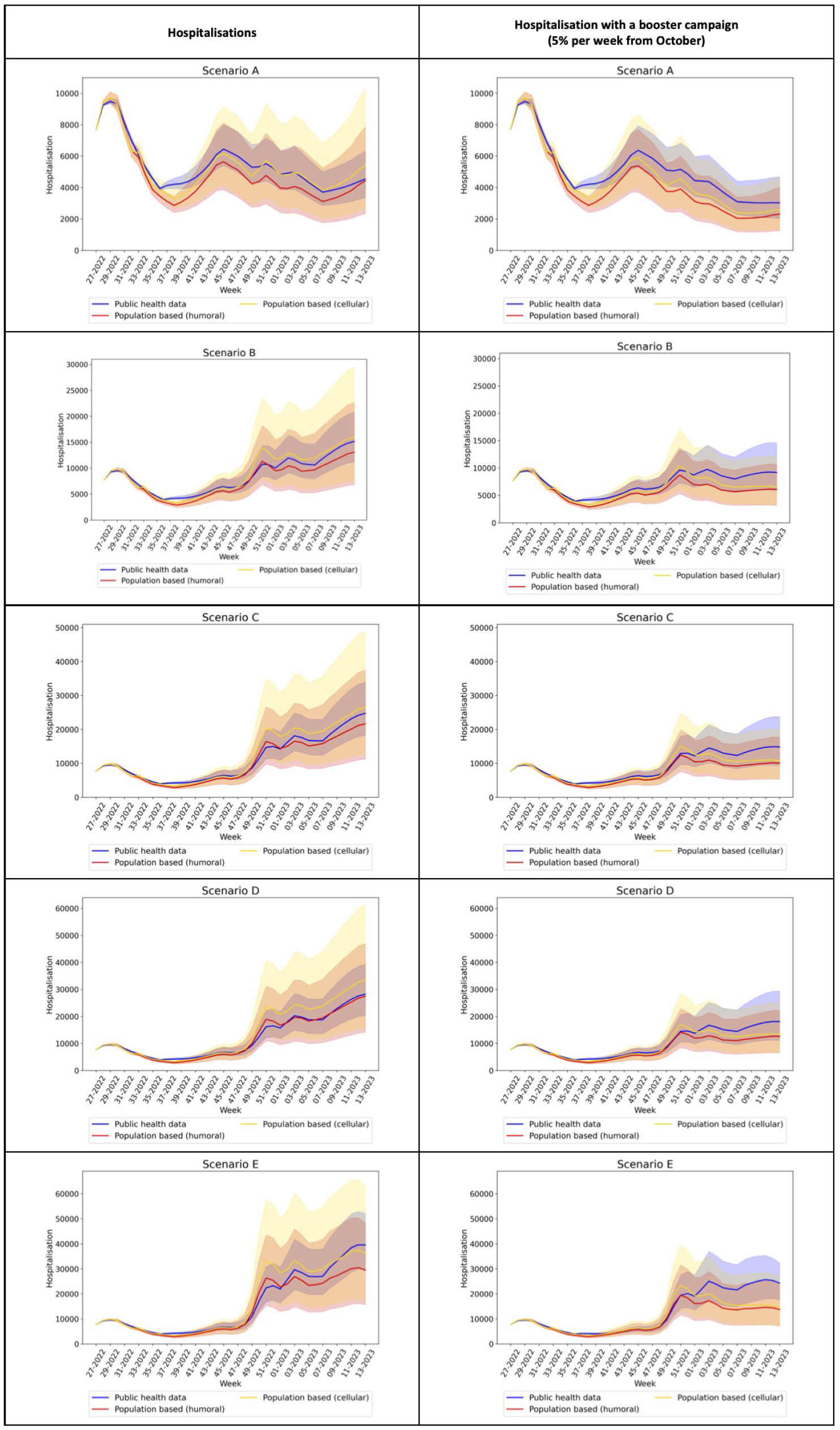
Scenarios A (Variant similar to BA.5 with vaccination), B (new variant with higher transmissibility), C (new variant with higher severity and transmissibility), D and E (new variant with higher transmissibility, severity and immune evasion) and associated hospitalization rates based on public health data, population based estimates (humoral and cellular) without (1) and with (2) booster campaigns in 2022/23 in Germany

In a base case scenario of variants with properties similar to BA.5, we found in all models peak hospitalizations for children and adults below 50% of those in winter 2022 during the BA1/2 wave in Germany (Table 4). A standard wild type vaccination campaign would not reduce hospitalizations by more than 20% as hospitalizations were predicted to mainly happen in the remaining weeks of 2022.

**Table 4.** Peak number of hospitalizations dependent on model used and scenario applied predicted for the winter 2022/23 in Germany

| Sce-<br>nario | Severity<br>in com-<br>parison<br>to BA5 | Im-<br>mune<br>eva-<br>sion | Trans-<br>missi-<br>bility | Peak ex-<br>pected be-<br>tween calen-<br>dar week a<br>and b | Peak hospitaliza-<br>tions expected kids<br>(0-4) compared to<br>BA 1/2 peak (460) | Peak hospitaliza-<br>tions expected kids<br>(5-14) compared to<br>BA 1/2 peak (440) | Peak hospitaliza-<br>tions expected<br>adults compared<br>to BA 1/2 peak<br>(12.000) | Peak hospital-<br>izations in +80<br>compared to<br>BA 1 / 2<br>(5000) | Reduction of hos-<br>pitalizations with<br>booster campaign<br>overall |
| --- | --- | --- | --- | --- | --- | --- | --- | --- | --- |
| Established model |  |  |  |  |  |  |  |  |  |
| Sce-<br>nario A | No new<br>variant | = | = | CW 42-50/22; | 60-70% | 20-40% | 30-40% | 40%-50% | 13% |
| Sce-<br>nario B | 1.3x | = | = | CW 1-7/23 | 100-120% | 20-40% | 100-130% | 120-150% | 25% |
| Sce-<br>nario C | 1.3x | 2x | = | CW 1-7/23 | 200-250% | 20-50% | 200-250% | 200-250% | 29% |
| Sce-<br>nario D | 1.3x | 2x | ++ | no | 220-270% | 30-40% | 230-290% | 230-290% | 30% |
| Sce-<br>nario E | 2x | 2x | ++ | yes | 270-300% | 30-60% | 300-340% | 300-400% | 33% |
| Population-based model using the combined endpoint with humoral immunity |  |  |  |  |  |  |  |  |  |
| Sce-<br>nario A | No new<br>variant | = | = | CW 42-50 | 20-40% | 10-20% | 20-70% | 20-60% | 9% |
| Sce-<br>nario B | 1.3x | = | = | CW 6-12/23 | 40-90% | 15-35% | 50-180% | 45-180% | 22% |
| Sce-<br>nario C | 1.3x | 2x | = | CW 6-12/23 | 100-180% | 30-60% | 100-300% | 100-300% | 27% |
| Sce-<br>nario D | 1.3x | 2x | ++ | CW 1-12/23 | 110-200% | 40-70% | 120-380% | 130-385% | 29% |
| Sce-<br>nario E | 2x | 2x | ++ | CW49-12/23 | 200-240% | 30-75% | 250-400% | 250-410% | 34% |
| Population model using combined endpoint with IGRA |  |  |  |  |  |  |  |  |  |
| Sce-<br>nario A | No new<br>variant | = | = | CW 42 –<br>51/22 | 20-40% | 10-20% | 20-80% | 30-75% | 10% |
| Sce-<br>nario B | 1.3x | = | = | CW 6 – 12/23 | 50-100% | 20-50% | 90-230% | 80-250% | 25% |
| Sce-<br>nario C | 1.3x | 2x | = | CW 6 – 12/23 | 100-200% | 30-80% | 120-380% | 120-400% | 31% |
| Sce-<br>nario D | 1.3x | 2x | ++ | CW 1 – 12/23 | 130-250% | 50-90% | 180-460% | 190-480% | 34% |
| Sce-<br>nario E | 2x | 2x | ++ | CW 49 –<br>12/23 | 190-300% | 40-100% | 260-500% | 260-550% | 33% |

CW = Calendar Week. There are 52 weeks in 2022 and 52 in 2023. All weeks are starting on Monday and ending on Sunday.

In a second scenario, with a new SARS-CoV-2 variant with 1.3 times the transmissibility of BA.5, but equal pathogenicity and immune evasion, we found a comparable height of peaks of hospitalizations at the beginning of 2023 in adults and children as seen during BA1/BA 2 in all three models; a vaccination campaign in our model was able to reduce overall hospitalizations up to 25% (Table 4).

In a third scenario with transmissibility increased as in scenario B but additionally increased pathogenicity, all models predicted a surpassing of peaks seen during the BA1/BA2 wave to slightly different degrees, and a reduction of overall hospitalizations with a vaccination campaign of about a third.

In scenario D and E, we found that even higher peaks of hospitalizations are possible in the models (up to 300% of peaks in BA1/2, both in adults, but also in children) if transmissibility or immune evasiveness of variants were relevantly raised. Vaccination and/or booster campaigns would reduce in these scenarios overall hospitalizations by up to 40% (Table 4).

Overall, the model based on data from the population panel predicted slightly lower and slower peaks in comparison to the model using public health surveillance data. In the third model integrating cellular immunity estimates from the IGRA measurements, peaks and the overall number of hospitalizations were increased in comparison to the model based on humoral immunity, but lower than in the model based on public health surveillance data

## Discussion

In this study, we showed how a rapid population based panel from June and July 2022 in Germany could be used to derive and validate proxies for protection from severe course of SARS-CoV-2 infection, and how this additional knowledge affected modelling studies aiming at an ad-hoc estimation of healthcare burden for the approaching winter.

More than 95% of the study population in most adult age groups has had more than three exposures to either the SARS-CoV-2 virus or vaccination with humoral immune correlates, respectively. 68.3% of those aged above 65 years and 45.9% of those over 80 years had not yet had a fourth vaccination. More than 90% of people in our sample were willing to be vaccinated or perhaps revaccinated in fall. When we assessed further booster vaccination campaigns, we found that these would be able to decrease the number of SARS-CoV-2 hospitalizations in the assessed scenarios by up to 40% if started in October at 5% of the population per week in particular in those scenarios with more pathogenic SARS-CoV-2 variants.

We found that overall more than 34% in all adult age groups had four exposures to SARS-CoV-2 (infection/vaccination) and confirmation of humoral immune correlates, with one of these exposures in 2022. We hypothesized that this group would have the highest protection against severe course of disease and at least some protection against re-infection based on available literature ^30^. However, we also found that even in this group 24.5% did not have interferon-gamma release after stimulation with spike specific antigens as detected by SARS-CoV-2 specific IGRA. The same was true for 36.7% of those who had had at least three exposures with confirmed humoral immunity. Positivity of IGRAs was age-dependent, with those over 80 years of age having lower odds of positivity. Overall, in our study positive IGRA was detected in 65.7 % of participants. Previous studies showed high sensitivity of IGRA to detect very recent infections, with 100% directly post-exposure and a decline to 79.5% after 10 months ^31^.

Assessing different scenarios of SARS-CoV-2 variants and vaccination strategies with three compartmental models including these population-based estimates, we calculated that hospitalization peaks for adults and children seen in the BA1/2 wave in January and February would not be surpassed without the introduction of a new variant or significant immune waning not foreseen in our models (Scenario A). Scenarios assessing hypothetical introductions of new SARS-CoV-2 variants with higher transmissibility, higher pathogenicity and higher immune evasiveness showed that it is likely possible to relevantly surpass previous hospitalization peaks seen in the winter waves 2020, 2021 and the BA1/2 wave. The results obtained here were in line with modelling studies from the time before the BA.5 summer wave in the UK where a higher and more detail amount of national population-based information was available ^29^. It was also in line with most models participating in a statement from the newly established central modelling network in Germany (MONID) in September 2022 ^18^, to which this group also reported.

While the overall prediction that no surpassing of clinical capacities would take place in Scenario A was correct, both our model and modelling estimates from the groups described above underestimated the height of the first hospitalization peak seen within a second BA.5 wave in calendar weeks 39 to 44 by around 30 - 40%. In contrast, the total number of hospitalizations (Supplementary Figure 3) ^35^ was not relevantly underestimated. In our opinion, this was both an effect of our model assuming a more prolonged second wave of the variant as well as assuming a similar age-specific distribution of infections compared to the first BA.5 wave. However, the actual second BA.5 wave had rather short but steep dynamics and a relevantly larger relative number of cases from the older age groups, resulting in an overall higher hospitalization risk per case during the second BA.5 wave. More accurate use of estimates of age-specific underdetection during the BA.5 summer wave might have helped to predict the larger contribution of elderly during the second BA.5 wave ^22^.

Our decision to use combined endpoints for protection against infection and severe disease progression based on the available literature is supported by the finding that there is a clear trend toward lower neutralizing activity in the lower categories of this endpoint than in the higher categories. For this endpoint, we designed the highest protection to be at least four exposures, one of which (vaccination or infection) occurred in 2022, which is also supported by a clear trend toward higher neutralizing activity among those with recent infections.

It is currently unclear how much complex model-usable endpoints of protection against severe course of disease and infection benefit from including neutralization titers or indirect correlates for cellular response like IGRAs. While IGRAs are a well-established tool for measuring the immune response to *Mycobacterium tuberculosis* ^26^, they represent a new tool for estimating specific cellular immune responses after SARS-CoV-2 infection or vaccination. IGRAs have shown clinical capacity for the assessment of specific T cell immunity immediately after vaccination ^27^, with the manufacturers reporting a sensitivity of 80% and specificity of 93% for detecting reported vaccination. This is especially true for patients with lack of humoral immunity, due to autoimmune disease or B-cell depleting therapy ^27,32^. Whether IGRA measurement in population-based studies can support the prediction of future risk of severe disease is still unclear ^31,33,34^.

The gap between modelling results based on population-based estimates and actual hospitalizations in week 39 - 44 could also be a result of underestimating the protection of the elderly against severe course of disease within this combined endpoint. In subsequent research, including prospective surveys and measurements, we will investigate whether the combined endpoint could be improved by assuming higher protection levels for those vaccinated with antibodies to nucleocapsid, by including a waning function by the time since last vaccination for those without antibodies to nucleocapsid, and by adding estimates from cellular immunity. Combined endpoints including cellular immunity might be more appropriate for older age groups where protection could then be assumed to be decreased due to lower IGRA positivity.

Our study has limitations. The population-based part is limited by not including children and typically underrepresented groups like asylum seekers and refugees. We were also regionally limited as we only included data from eight (though exemplary) regions in Germany. The used ODE models are limited by not accounting for underdetection estimates in the population or incidental hospitalizations during the BA.1/2 wave.

Despite these limitations, we show that rapid, adaptive population panels using combined endpoints can work to directly parametrize scenario and forecast models during epidemics in Germany. The data from our study and other studies were made available in aggregate format ^17^ to a central modelling platform ^18^ currently being established in Germany so that other modelling groups can use it to parametrize their models.

## Conclusion

Although we show protection in the majority of the population against severe course of disease measured by at least three exposures or vaccination and confirmed by humoral immune correlates even quite moderate changes in transmissibility or pathogenicity of new SARS-CoV-2 variants could lead to relevant hospital burden surpassing previous waves if no remedial action is taken. Future epidemic panels should prospectively evaluate complex endpoints for protection against SARS-CoV-2 infection and severe course of disease.

## Data Availability

All data produced in the present study are available upon reasonable request to the authors. The anonymized data for this study will be made available to other academic researchers.
Institutions can apply for the data via.

https://www.rki.de/DE/Content/InfAZ/N/Neuartiges_Coronavirus/Daten/Klinische_Aspekte.xlsx?__blob=publicationFile

https://zenodo.org/record/7177592#.Y4okYnbMI2w)

## Additional Information

Table S1: Protection against severe course of disease and infection for different combinations of vaccination and infection in adults with the Omicron variant

Table S2: Combined endpoint in those with and without IGRA Age distribution of the MuSPAD participants with blood samples grouped by combined endpoints including IGRA test results

Table S3: Parametrization of the combined endpoints with and without humoral immune correlations as well as positive or negative IGRA based on literature synthesis

Figure S1: Scatterplot of correlation between anti-S by two variants Wu01 and BA.5 (A, B) and anti-NC antibody titers (C, D) based on Roche ELISA test results and neutralization.

Figure S2: Boxplot of BA.5 neutralizing antibody response by time since last SARS CoV-2 vaccination and last SARS-CoV-2 infection stratified by IGRA

Figure S3: Weekly number of COVID 19 cases hospitalized, cumulated over all age groups in Germany

## Acknowledgements

We appreciate all MuSPAD participants supporting and endorsing our study. We are grateful to our colleagues in Oldenburg (especially Heike Adam) and Cologne for the laboratory analyses. We thank BOS112 (Tim Balz), NAKO (in particular Sabrina Sistig) and Barbora Kessel (former HZI) for collaborating with us. We thank IPSOS for great technical appointment management and HUB MHH for the sample bio banking.

## Data sharing statement

The anonymized data for this study will be made available to other academic researchers. The minimal dataset includes study site information, assay information, sample type, demographic information, self-administered diagnostic anamneses and lab results (NC, S spike, IGRA and NAb). For further details contact. Institutions can apply for the data via.

## Informed Consent Statement

Prior to study participation, all participants provided signed informed consent.

## Conflicts of Interest

The authors declare no conflict of interest.

### Author Contribution

Conceptualization: A.K., B.L.;

Data quality: M.J.H., J.O., V.J.,MA.K.;

Formal analyses: M.H., V.J, I.R., M.J.H, B.L., A.K. M.D., I.v.H., M.B;

Visualization: M.H., V.J., I.R;

Project administration: C.K., D.G., M.H., M.S.;

Laboratory analysis: A.P., F.K., M.S, A.D., N.S., M.S.;

Evidence synthesis: B.L., M.D., I.v.H., M.B., O.H.

Writing-original draft: M.H., V.J., B.L., A.K., I.R.;

Funding acquisition: B.L., A. K., G.K.;

Writing-review & editing: M.S., M.J.H., J.O., V.J., MA.K, C.K., D.G., M.H., B.L., A.K., F.K., M.S., I.R., A.P., A.D.,N.S.,O.H.,G.K. M.D.,I.v.H., M.B

All authors have read and agreed to the published version of the manuscript.

