## Supplemental Data 1 for "Bridging the gap – estimation of 2022/2023 SARS-CoV-2 healthcare burden in Germany based on multidimensional data from a rapid epidemic panel"

### Supporting Information

MuSPAD sampling/survey period:

The blood sampling period took place from June 6th, 2022 to July 3rd, 2022. We chose three existing MuSPAD study sites: City Region Aachen (around 560,000 inhabitants), Magdeburg (around 236,000 inhabitants) and Hannover (around 536,000 inhabitants). From June 2022 on, we send out our online and paper-based survey to all established MuSPAD participants (Freiburg, Reutlingen, Chemnitz, Greifswald, Magdeburg, Hannover, Aachen, and Osnabrück).

Lab analysis:

The serum samples obtained were analyzed centrally at the Institutes of Clinical Chemistry and Laboratory Medicine in Oldenburg and Greifswald. The Elecsys® Anti-SARS-CoV-2 S and Elecsys® Anti-SARS-CoV-2 NC (both Roche Diagnostics, Mannheim, Germany) were used for the determination of the quantitative antibody response against the S and N antigen of SARS-CoV-2. For Nucleocapsid (NC) protein we used ≥0.8 BAU/ml cut-offs for seropositivity. We also performed SARS-CoV-2 specific interferon-gamma release response (IGRA) using SARS-CoV-2 LIASION QuantiFERON-TB Gold Plus Qiagen®. For the blood collection, we used Lithium-Heparin Monovette and transferred them into specialized QFT-Plus Blood Collection with 24 h. We incubated the samples for 16 to 24 hours at 37°C, centrifuged the tubes at 2000 to 3000 RZB (g) for 15 minutes. Subsequently we detected released IFNγ in harvested plasma by ELISA.

For the neutralization assay, we investigate the activity of heat-inactivated (56°C) serum against lentivirus (HIV)-based pseudoviruses. Our assay is a further development of the protocol of Crowford et al. [25]. In the methodology used, it was first described by us in the paper by Vanshylla et al.[24]. For our serum samples, the 50% inhibitory serum dilution (ID50) was tested against pseudoviruses with the spike protein of the Wu01 variant and the BA.4/5 variant. Samples were tested in a 1:3 dilution series at dilutions ranging from 1:10 to 1:21,870 - this is within the quantification range of the assay.

**Supplementary Table S1:** Protection against severe course of disease and infection in adults for different combinations of vaccination and infection with the Omicron variant[1]

| **Exposures** | **VE**[2] | **VE in studies without information on follow-up** | **VE 0-13 days** | **VE 14-90 days** | **VE 91-180 days** | **VE >180 days** |
|---|---|---|---|---|---|---|
| 4 Exposures | Infection[3] | 46-96% (n=6) | 36% (n=1) | 88-98% (n=2) | 72-96% (n=2) | 82% (n=1) |
| | Severe course of disease[4] | 68-97% (n=4) | | | | |
| | Death | 81% (n=1) | | | | |
| 3 Exposures | Infection[3] | 35-78% (n=13) | 18-69% (n=8) | 32-89% (n=14) | 5-96% (n=10) | 34-68% (n=3) |
| | Severe course of disease[4] | 34-99%, AZ 30% (n=9) | 65-95% (n=3) | 59-97% (n=10) | 52-95% (n=10) | 65% (n=1) |
| | Death | 66-98% (n=2) | 71-86% (n=1) | 82-86% (n=2) | 87% (n=1) | 77% (n=1) |
| 1-2 Exposures | Infection[3] | -6-90% (n=17) | 10-23% (n=2) | 12-82% (n=16) | 3-70% (n=15) | -13-56% (n=9) |
| | Severe course of disease[4] | 41-97% (n=10) | 28-46% (n=1) | 42-96% (n=10) | 22-91% (n=14) | 12-88% (n=6) |
| | Death | 66-90% (n=3) | -9-29% (n=2) | 60-62% (n=2) | 57-70% (n=1) | 49-57% (n=2) |

[1] Interims analysis (see https://zenodo.org/record/7177592#.Y4okYnbMI2w) based on 27 identified studies from View hub on 9/27/2022; International Vaccine Access Center, Johns Hopkins Bloomberg School of Public Health, World Health Organization, Coalition for Epidemic Preparedness Innovations. Results of COVID-19 Vaccine Effectiveness Studies: An Ongoing Systematic Review: Weekly Summary Tables. Updated September 22, 2022. Available from: URL: https://view-hub.org/sites/default/files/2022-09/COVID19_Vaccine_Effectiveness_Transmission_Studies_Summary_Tables_20220922.pdf.

[2] VE Vaccine effectiveness (reference: no vaccination, infection, or antibodies); range of point estimates of the included studies

[3] Definitions included every, symptomatic, or documented infection

[4] Definitions included hospitalization, admission on intensive care unit, and/or death

**Supplementary Table S2:** Combined endpoint in those with and without IGRA Age distribution of the MuSPAD participants with blood samples grouped by combined endpoints including IGRA test results

| Age groups in years | 18-34 n (%) | 35-49 n (%) | 50-64 n (%) | 65-79 n (%) | 80+ n (%) | Overall n (%)[1] |
|---|---|---|---|---|---|---|
| **Combined endpoints in those with blood sampling n=2898** | | | | | | |
| 3 exposures + one in 2022 | 85 (31.1%) | 168 (35.1%) | 297 (26.0%) | 372 (42.7%) | 69 (51.5%) | 991 (34.2%) |
| 3 exposures | 170 (62.3%) | 275 (57.5%) | 786 (68.9%) | 481 (55.2%) | 63 (47.0%) | 1775 (61.2%) |
| 1-3 exposures, or no immune correlates | 18 (6.6%) | 34 (7.1%) | 51 (4.5%) | 15 (1.7%) | 1 (0.7%) | 119 (4.1%) |
| 0 exposures, no immune correlates | 0 (0%) | 1 (0.2%) | 7 (0.6%) | 4 (0.5%) | 1 (0.7%) | 13 (0.4%) |
| Total | 273 | 478 | 1141 | 872 | 134 | 2898 |
| **Combined endpoint including IGRA result in those with IGRA* sampling n=973 [1,2]** | | | | | | |
| 3 exposures + one in 2022 (n; %IGRA negative) | 33; 27.0% (5; 2%) | 58; 31.2%; (5; 8.6%) | 60; 19.5% (9; 15.0%) | 101; 36.9% (31; 30.7%) | 38; 23.6% (21; 55.3%) | 290, 29.8% (71; 24.5%) |
| 3 exposures (n; % IGRA negative) | 81; 66.4% (21; 25.9%) | 103; 55.4%; (24; 23.3%) | 224; 73.0% (68; 30.4%) | 168; 61.3% (90; 53.6%) | 45; 28.0% (25; 55.6%) | 621; 63.8% (228; 36.7%) |
| 1-3 exposures, or no immune correlates (n; % IGRA negative) | 8; 6.6% (6; 75.0%) | 24; 12.9% (12; 50.0%) | 21; 6.8% (8; 38.1%) | 3; 1.1% (1; 33.3%) | 0 | 56; 5.8% (27; 48.2%) |
| 0 exposures, no immune correlates (n; % IGRA negative) | 0 | 1;0.5% (1; 100%) | 2; 0.7% (2; 100%) | 2; 0.7% (2; 100%) | 1; 0.6% (100%) | 6; 0.6% (6; 100%) |
| Total | 122 | 186 | 307 | 274 | 84 | 973 |

*IGRA= interferon-gamma release response; [1]43 participants did not specify age; [2]62 participants did not indicate a combined endpoint

**Supplementary Table S3:** Parametrization of the combined endpoints with and without humoral immune correlations as well as positive or negative IGRA based on literature synthesis

| Groups of combined endpoints | **Vaccination/Infection/Immunity** | **Parametrization of models** | Protection vs symptomatic infection for fully protected | Protection vs hospitalization for fully protected | Protection vs ICU* for fully protected |
|---|---|---|---|---|---|
| Group I<br>3 + 1 exposure in 2022 | 3 exposures (vaccination or infection) with humoral immune correlates* + one infection/vaccination in 2022 | Probably high protection vs severe course of disease<br>Some protection vs infection | 40-60% | 70-90% | 70-90% |
| Group II<br>3 exposures | 3 exposures (vaccination or infection) with humoral immune correlates* | Moderate protection vs severe course of disease<br>Some protection vs infection | 10-30% | 50-70% | 50-70% |
| Group III<br>1-3 exposures or no immune correlates | 3 or more exposures without immune correlate, 1-2 exposures with or without immune correlate or 0 exposures with at least 1 positive humoral immune correlate* | Some protection vs severe course of disease<br>Little protection vs infection | 0 | 20-40% | 20-40% |

*ICU= intensive care unit

**Supplementary Figure S1:** Scatterplot of correlation between anti-S by two variants Wu01 and BA.5 (A, B) and anti-NC antibody titers (C, D) based on ELISA test and neutralization

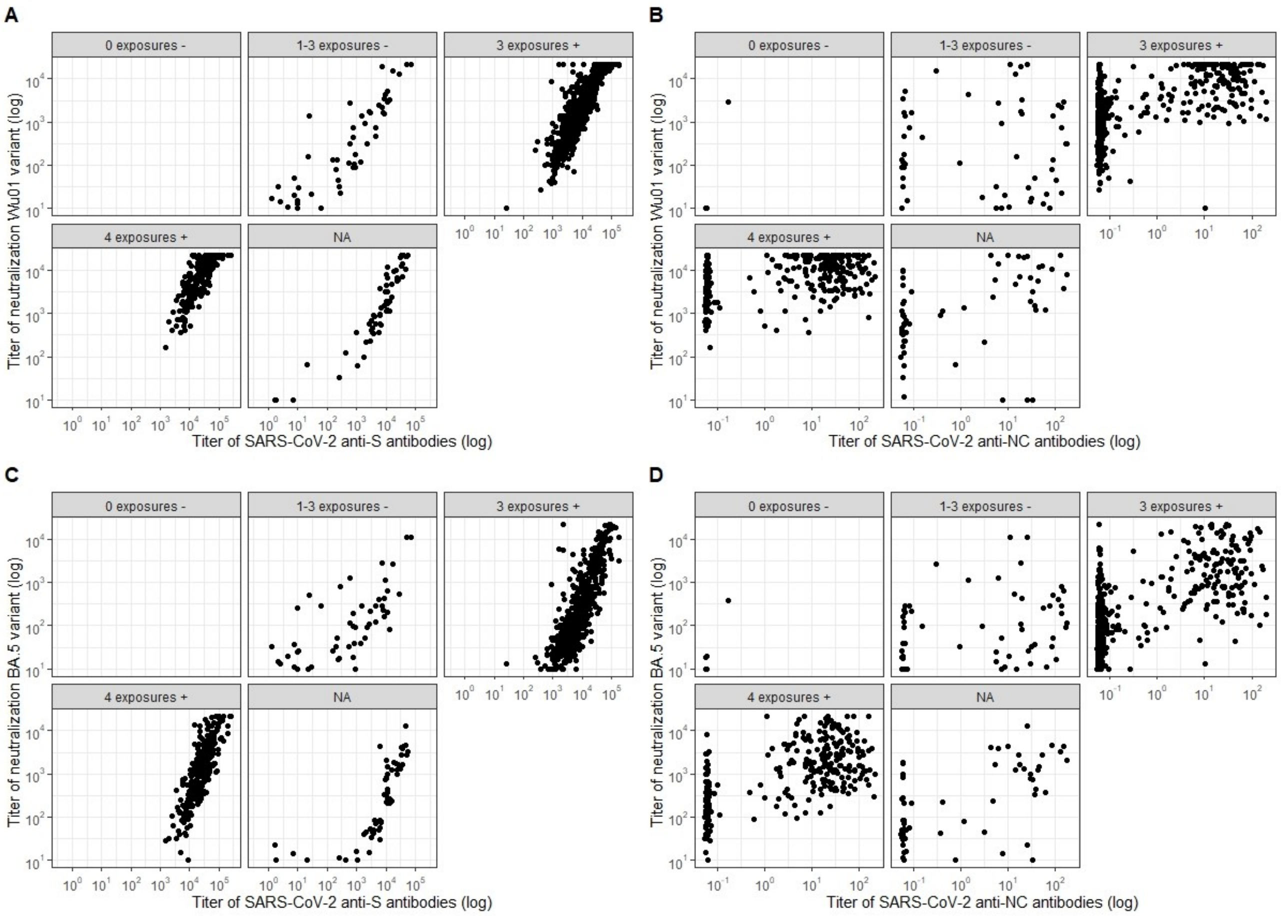

**Supplementary Figure S2:** Boxplot of BA.5 neutralizing antibody response by time since last SARS CoV-2 vaccination **(a)** and last SARS-CoV-2 infection **(b)** stratified by IGRA

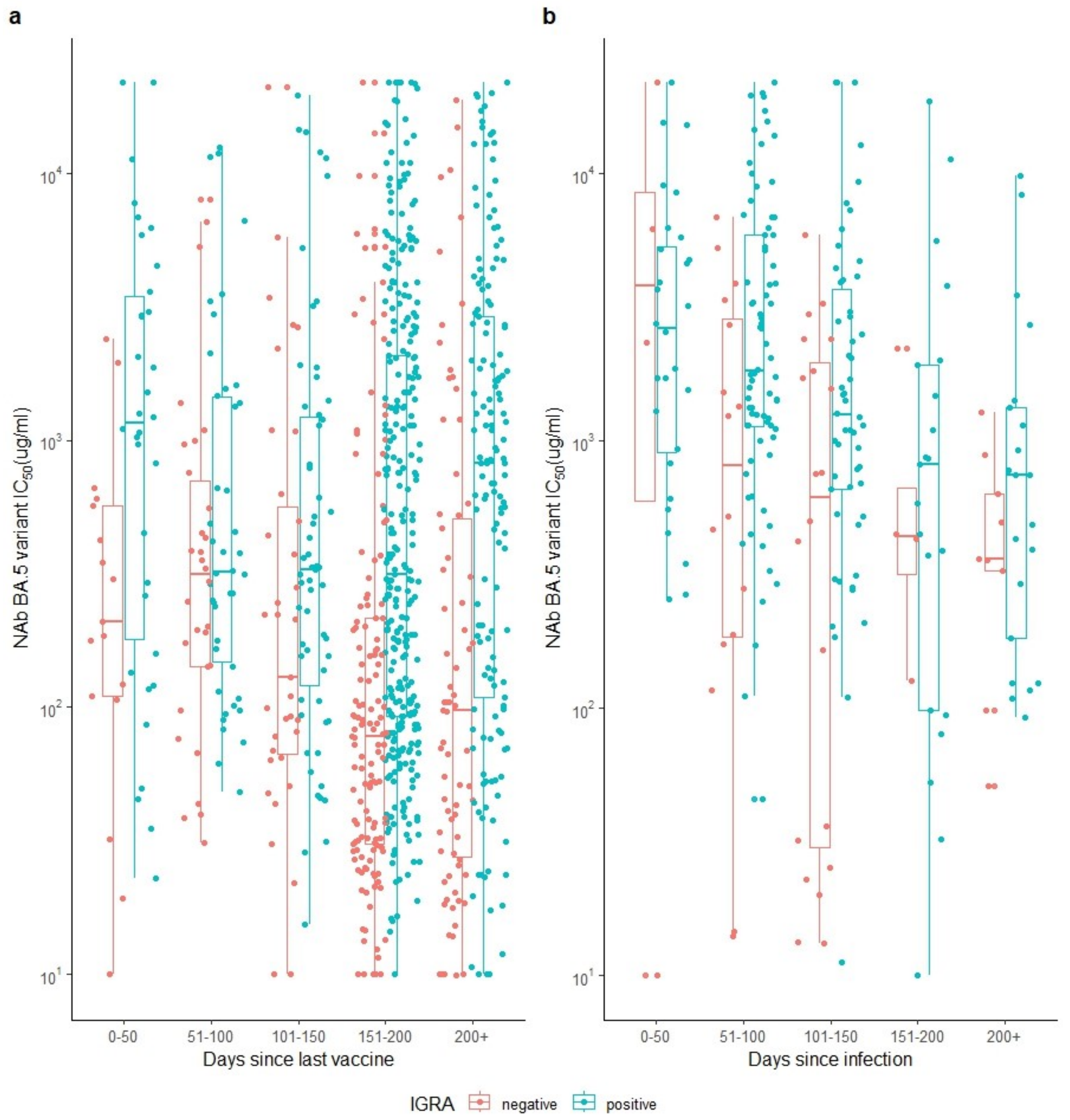

**Supplementary Figure 3:** Weekly number of COVID 19 cases hospitalized, cumulated over all age groups in Germany (https://www.rki.de/DE/Content/InfAZ/N/Neuartiges_Coronavirus/Daten/Klinische_Aspekte.xlsx?__blob=publicationFile

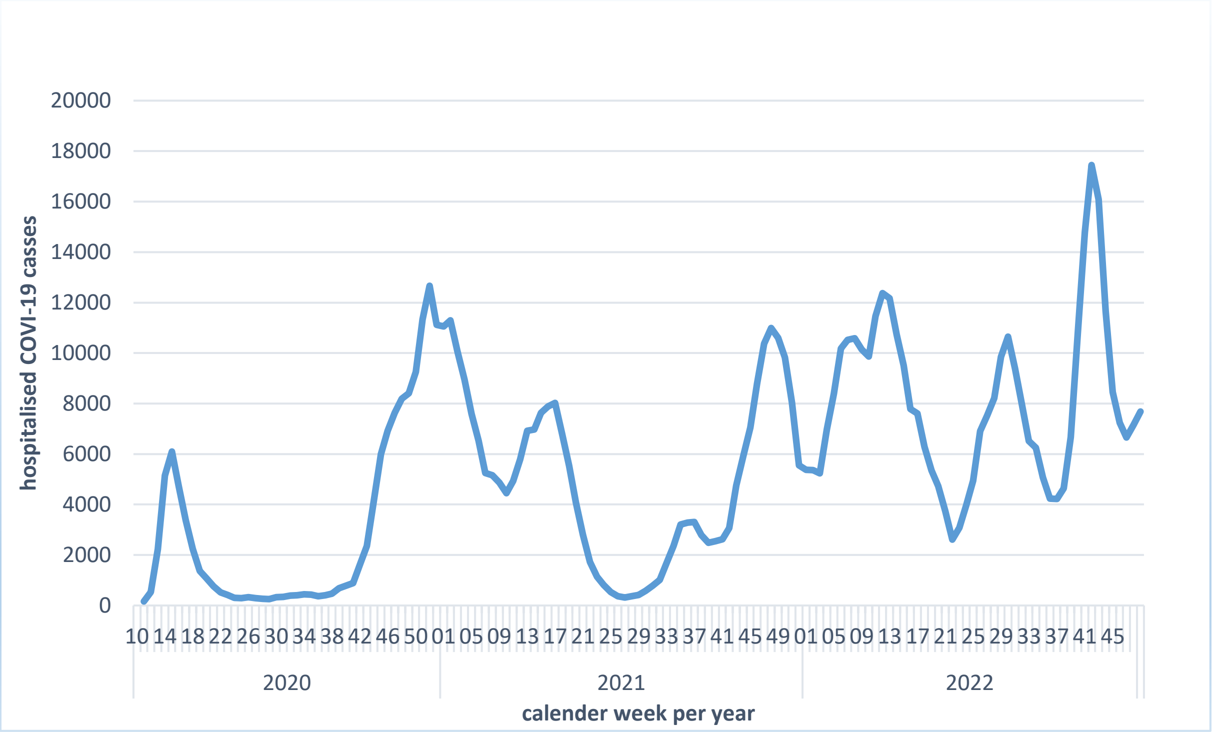
